# Evaluation of a surrogate virus neutralization test for high-throughput serosurveillance of SARS-CoV-2

**DOI:** 10.1101/2021.02.24.21252047

**Authors:** Joachim Mariën, Johan Michiels, Leo Heyndrickx, Antoine Nkuba-Ndaye, Ann Ceulemans, Koen Bartholomeeusen, Joule Madinga, Placide Mbala-Kingebeni, Veerle Vanlerberghe, Steve Ahuka-Mundeke, Lin-Fa Wang, Kevin K. Ariën

## Abstract

High-throughput serological tests that can detect neutralizing antibodies against SARS-CoV-2 are desirable for serosurveillance and vaccine efficacy evaluation. Although the conventional neutralization test (cVNT) remains the gold standard to confirm the presence of neutralizing antibodies in sera, the test is too labour-intensive for massive screening programs and less reproducible as live virus and cell culture is involved. Here, we performed an independent evaluation of a commercially available surrogate virus neutralization test (sVNT, GenScript cPass^™^) that can be done without biosafety level 3 containment in less than 2 hours. When using the cVNT and a Luminex multiplex immunoassay (MIA) as reference, the sVNT obtained a sensitivity of 94% (CI 90-96%) on a panel of 317 immune sera that were obtained from hospitalized and mild COVID-19 cases from Belgium and a sensitivity of 89% (CI 81-93%) on a panel of 184 healthcare workers from the Democratic Republic of Congo. We also found strong antibody titer correlations (r_s_>0.8) among the different techniques used. In conclusion, our evaluation suggests that the sVNT could be a powerful tool to monitor/detect neutralising antibodies in cohort and population studies. The technique could be especially useful for vaccine evaluation studies in sub-Saharan Africa where the basic infrastructure to perform cVNTs is lacking.

## Introduction

One year after the emergence of severe acute respiratory syndrome coronavirus 2 (SARS-CoV-2) in China, more than 2 million fatal and 100 million diagnosed cases are recorded worldwide (Johns Hopkins University, 2020). Serosurveillance data suggests many more undiagnosed cases as national rates typically range between 5-15% with local rates up to 50%(1). Accurate serological data will also play a crucial role during the next phase of the pandemic. Indeed, massive vaccination campaigns are currently in progress and their efficacy needs to be monitored continuously to adjust control and prevention policies. Furthermore, while it is known that most people develop a long-lasting antibody immunity (at least 6 months after infection)(2,3), the sporadic detection of reinfections in immunocompetent individuals(4) and the emergence of new variants that might evade the antibody response(5) highlight the need to better understand SARS-CoV-2 antibody immunity at an individual level by directly determining the neutralizing antibody (NAb) level rather than just total binding antibodies (Babs).

While a plethora of serological tests became available months after the emergence of SARS-CoV-2, not all of them are appropriate for large-scale serosurveillance. Most high-throughput tests detect total BAbs only, such as enzyme-linked immunosorbent (ELISA), lateral flow (LFA) or multiplex (MIA) immunoassays (6). These tests can be run in basic diagnostic labs and are mainly used to confirm past infection, but are unable to directly show the presence of neutralizing antibodies in serum. The latter are typically detected with conventional virus neutralization tests (cVNT) which are labour-intensive and take 4-5 days to complete by highly trained staff in a BSL3 laboratory. Showing the direct presence and determining the level and longevity of NAbs will be crucial for vaccine evaluation or serosurveillance in populations where cross-reactivity of BAbs against other related coronaviruses is likely, such as in sub-Saharan Africa (7). Furthermore, detecting NAbs might be the only way to show past SARS-CoV-2 infections in particular wildlife populations for which secondary antibodies are unavailable (e.g. in populations of bat, pangolin or mink (8,9)). To overcome the difficulties of the cVNT, a surrogate viral neutralization test (sVNT) was recently developed that can be completed in 1-2 hours in a BSL2 laboratory and made commercially available by GenScript (10). The test uses the principle of an ELISA to measure the neutralizing capacity of anti-SARS-CoV-2 antibodies by inhibiting the interactions between the receptor-binding domain (RBD) of the spike protein and ACE2 cell receptors, mimicking the virus’ neutralization process. Here, we performed an independent evaluation of this commercial sVNT on sera from COVID-19 cases that were screened on NAbs by cVNT and BAbs by MIA in our lab.

## Methods

Serum samples from Belgium (n=316) and the Democratic Republic of Congo (n=184) were used to assess the sVNT (supplementary data file 1). Our Belgian panel consisted of serum samples that were obtained at different time intervals after PCR confirmation: 163 samples were taken 1-5 weeks (recent), 45 samples 6-20 weeks (intermediate) and 108 samples 20-24 weeks (old) after PCR confirmation (6,11). All serum samples were obtained in the period between March and August 2020 in different hospitals in Belgium and were from either hospitalized COVID-19 patients (severe illness, *n*=144) or health-care workers (mild or asymptomatically infected, *n*=172). Of these, 60 hospitalized cases were sampled two or three times over a period of one or two weeks, while all other samples belonged to different individuals. Since we (and many others) showed that SARS-CoV-2 antibody titers decrease significantly after the initial increase and depend on disease illness (6), this diverse panel assured that both high and low antibody titer sera were included. All Congolese samples were obtained in July-August 2020 during a cross-sectional survey of staff working in healthcare facilities in Kinshasa, the capital of the DRC. As we did not know the infection status of these participants (no PCR tests were performed), this panel included serum from people having been exposed or not to SARS-CoV-2 since the start of the pandemic in March 2020. Neutralizing antibody titers were only assessed in Congolese samples that were suggested to be positive by one of the following Bab-assays: Euroimmun Anti-SARS-CoV-2-spike IgG, Quickzen (Zentech, Belgium) IgG and IgM(12). All serum samples were inactivated by heating at 56°C for 30 min.

The surrogate virus neutralisation test (sVNT) (GenScript cPass^™^, USA, L00847) was performed according to the manufacturer’s instructions(10). Samples, positive and negative controls were diluted 1:10 with sample dilution buffer. The dilutions were mixed with horseradish peroxidase conjugated recombinant SARS-CoV-2 RBD solution and incubated for 30min at 37°C. The mixtures were subsequently incubated for 15 minutes at 37°C in a capture plate that was pre-coated with hACE2 protein. After a washing step, tetramethylbenzidine (TMB) solution was added and the plate was incubated in the dark at room temperature for 15min. Stop solution was added to quench the reaction and the absorbance was immediately read at 450nm on a ELISA microplate reader. The percentage inhibition was calculated as 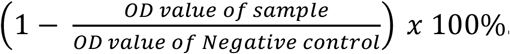. Sensitivity was calculated at a low (at 20%) and high (at 30%) inhibition cut-offs, which corresponds to a specificity of 98 and 100% respectively(10).

Samples were also screened by an in-house cVNT and Luminex MIA as reference, which are described in detail in Mariën et al 2020 (6). Briefly, for the cVNT, serial dilutions of serum (1/50-1/1600) were incubated with 3xTCID100 of a primary isolate of SARS-CoV-2 during 1 h. This solution was added to Vero cells (18.000cells/well) in a 96well plate and incubated for 5 days (37 °C / 7 % CO2). The Reed-Muench method was used to calculate the neutralising antibody titre that reduced the number of infected wells by 50 % (cVNT_50_) or 90 % (cVNT_90_). Samples were still considered to be positive if more than 10% reactivity was observed at a 1/50 serum dilution. For the Luminex MIA, recombinant receptor binding domain (RBD) and Nucleocapsid protein (NCP) (BIOCONNECT, The Netherlands) were coupled to 1.25×10^6^ paramagnetic MAGPLEX COOH-microspheres from Luminex Corporation (Texas, USA). After incubation of beads and diluted sera (1/300), a biotin-labelled anti-human IgG (1:125) and streptavidin-R-phycoerythrin (1:1000) conjugate was added. Beads were read using a Luminex® Bio-Plex 100/200 analyzer. Samples were considered to be positive if the fluorescent signal >2x standard deviation + mean of negative controls (*n*=96) for both antigens, which corresponds to a specificity of 99% (6). Results were expressed as signal-to-noise ratios. Only 198 samples were screened on IgG BAbs using the Luminex MIA.

## Results

From the 316 samples obtained from Belgian cases that tested PCR positive, 17 samples were seronegative for both VNTs and excluded from the sensitivity panel. While 12 of these samples were obtained <14 days after the PCR result (probably just before seroconversion started), five samples were taken 3-5 months after the PCR result. These five individuals might have been included in the panel on the basis of a false-positive PCR test or they seroreverted before serum sampling. Six samples that were negative in the cVNT_50_, but positive in the Luminex MIA (*n*=3) or the sVNT_20%_ (*n*=3), remained in the panel. Based on this final panel, we found that the sensitivity of the sVNT_20%_ was only slightly lower than the sensitivity of the cVNT_50_ (Δ4%)or the Luminex MIA (Δ3%) (**Table 1**). As expected, the sensitivity decreased at high specificity targets for the sVNT_30%_ (Δ9%) and the cVNT_90_ (Δ29%). The group of samples (n=16) that were negative on the sVNT_20%_ but positive on the cVNT_50_ mainly consisted of sera obtained from patients <14 days after the PCR result (n=4) or five months after infections (n=10). For most of these samples, less than 50% (but more than 10%) inhibition was observed at a 1/50 dilution in the cVNT_50_.

**Table 1:**
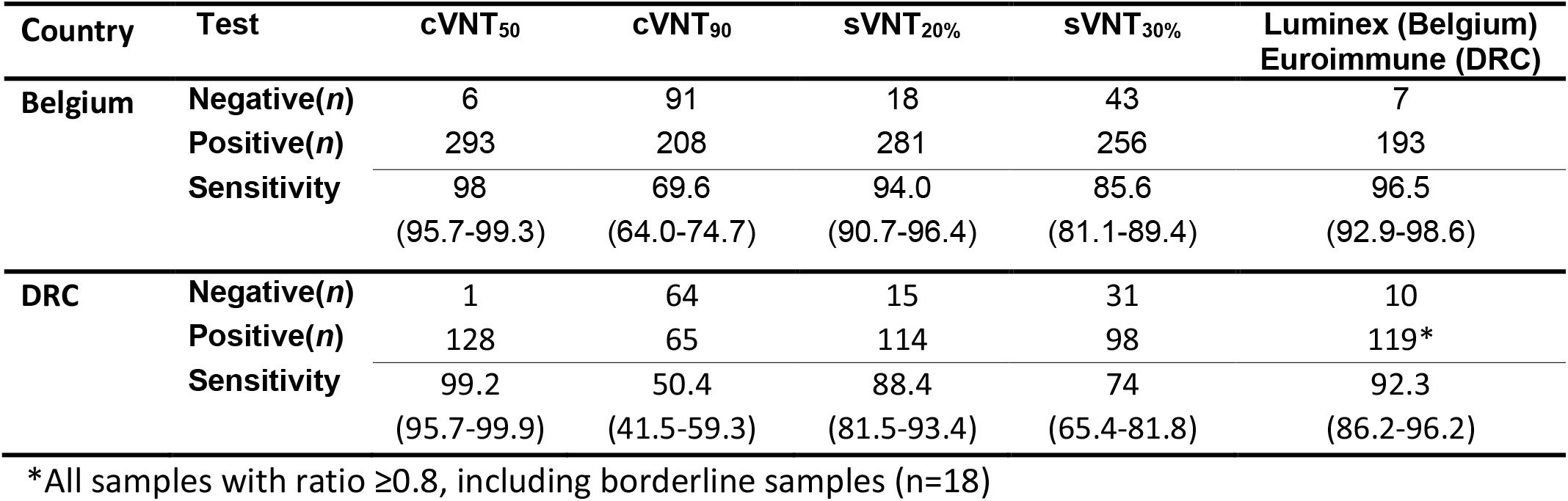
Sensitivity of the different serological tests and cut-offs used based on the final serum panels. Values between brackets represent 95% confidence intervals on the estimations.

| Country | Test | cVNT<sub>50</sub> | cVNT<sub>90</sub> | sVNT<sub>20%</sub> | sVNT<sub>30%</sub> | Luminex (Belgium)<br>Euroimmune (DRC) |
| --- | --- | --- | --- | --- | --- | --- |
| Belgium | Negative($n$) | 6 | 91 | 18 | 43 | 7 |
| | Positive($n$) | 293 | 208 | 281 | 256 | 193 |
|  | Sensitivity | 98 | 69.6 | 94.0 | 85.6 | 96.5 |
|  |  | (95.7-99.3) | (64.0-74.7) | (90.7-96.4) | (81.1-89.4) | (92.9-98.6) |
| DRC | Negative($n$) | 1 | 64 | 15 | 31 | 10 |
| | Positive($n$) | 128 | 65 | 114 | 98 | 119* |
|  | Sensitivity | 99.2 | 50.4 | 88.4 | 74 | 92.3 |
|  |  | (95.7-99.9) | (41.5-59.3) | (81.5-93.4) | (65.4-81.8) | (86.2-96.2) |

\*All samples with ratio $\geq 0.8$ , including borderline samples ( $n=18$ )

From the 184 samples obtained from Congolese participants with a positive commercial serology test, 55 tested negative on both the cVNT and sVNT and can be considered true negatives. One sample tested negative on the cVNT and positive on the sVNT (45% inhibition), which could be a potential false-positive result on the sVNT. If the sample is indeed false-positive (we cannot rule-out the possibility of a false-negative on the cVNT), the specificity of the sVNT (relative to the cVNT) would be 98.2% (CI 90.4-99.9) based on the Congolese panel for both the 20% and 30% cut-offs. Similar as for the Belgian samples, we found that the sensitivity of the sVNT_20%_ was lower (Δ11%) than the sensitivity of the cVNT_50_ (**Table 1**).

To test if we can use the sVNT as a high-throughput alternative for the more labor-intensive cVNT, we calculated correlations (r_s_) between the antibody titer proxies using the nonparametric Spearman rank test (R.3.6.1. statistical software). We found strong correlations (r_s_=0.85, p>0.0001) between the inhibition percentage of the sVNT and the dilution factors of the cVNT_50_ and cVNT_90_ (**Fig 1**). A strong correlation (rs=0.83, p>0.0001) was also observed between the inhibition percentage of the sVNT and signal-to-noise ratios of the RBD on the Luminex MIA, but not for the NCP (r_s_=0.44, p>0.0001) (**Fig 1**). The latter result is explained by the fact that the sVNT specifically detects antibodies that neutralize the RBD-ACE2 interaction, while antibodies directed against the NCP protein are most likely not relevant in this assay and less-neutralising in general.

**Fig 1.**
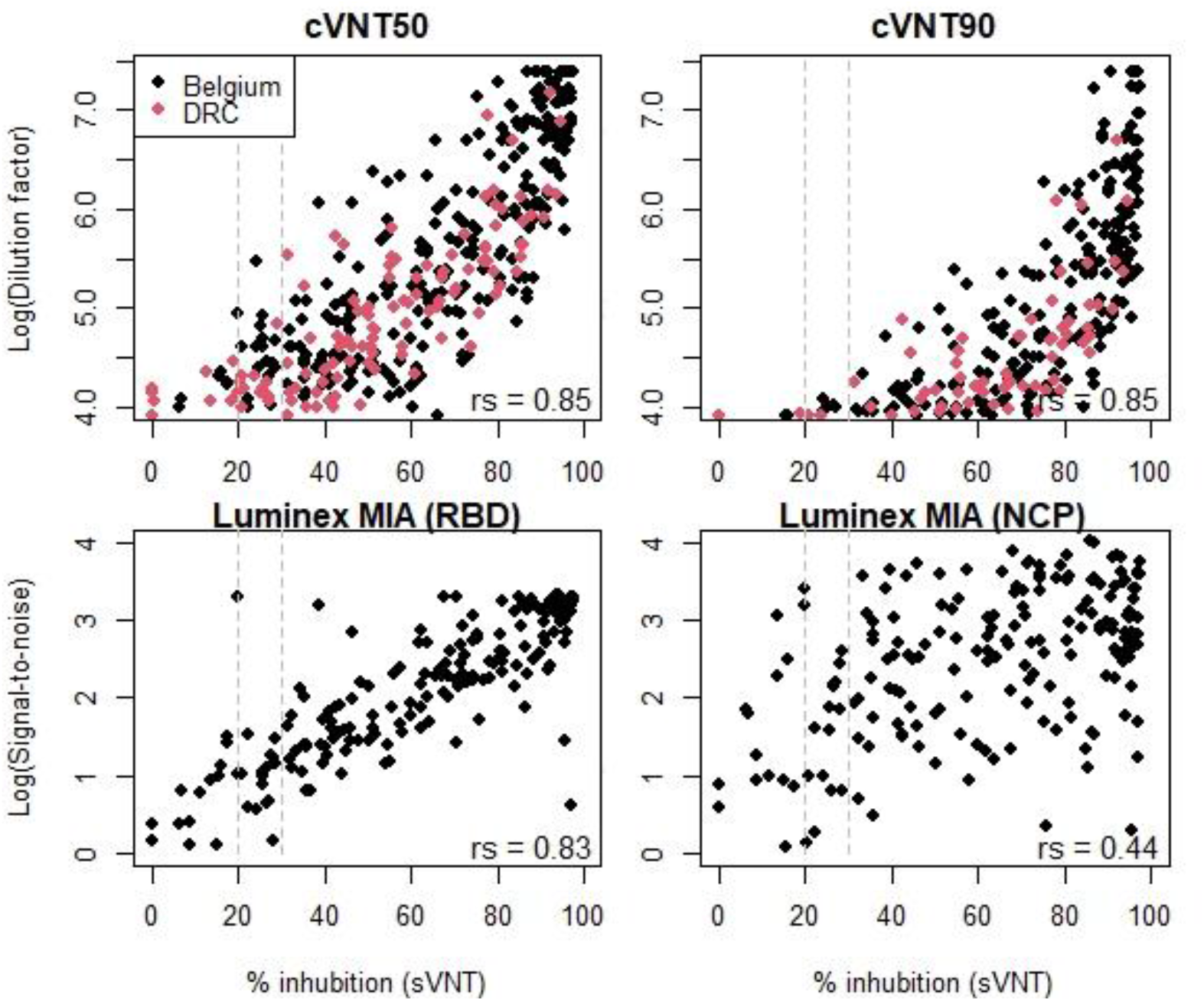
Correlations between the percentage of inhibition measured by the surrogate viral neutralisation test (sVNT) and the log(dilution factors or signal-to-noise ratio) for the conventional viral neutralization test (cVNT) or the Luminex multiplex immunological assay (MIA) as calculated by the nonparametric Spearman correlation test (r_s_). Seropositivity cut-off levels for the sVNT are indicated by the dashed grey lines at 20 or 30% inhibition. Negative samples on the cVNT or MIA were not included in these figures.

## Discussion

We found overall high concordance between the sVNT, the cVNT and the Luminex MIA in terms of sensitivity and antibody titer correlations. The sensitivity estimations of the sVNT_20_ in our study are in line with Bond et al(13) who evaluated the sVNT_20_ on serum from COVID-19 cases obtained 2-6 weeks after diagnosis in Australia. In contrast, Meyer et al(14) found a significantly lower sensitivity (83%) on serum from COVID-19 cases >14 days after symptom onset in the Netherlands. The latter also reports a significant difference in sensitivity between the two centres where their study was conducted, potentially explained by the underrepresentation of samples taken at later time points at one of the sites. This highlights again that the timing of serum collection relative to onset of disease can affect the performance characteristics for COVID-19 serological assays. A limitation of our study is that we only included a small panel of African samples (*n*=55) to assess the specificity of the test and we cannot rule-out cross-reactivity at low titers on the cVNT, which we used as reference. However, there is overall agreement that the specificity of the sVNT is acceptably high (94-99% at sVNT_20_ and 99-100% at sVNT_30_) when evaluated on a panel containing challenging samples, including other coronaviruses or other acute infections (cross-reactivity against SARS-CoV-1 is noted)(8,10,13,14).

In conclusion, our results suggest that the commercial sVNT could be a powerful tool to determine neutralising antibodies in cohort and population studies, although other high-throughput assays (such as a Luminex MIA) might outperform the sVNT in terms of individual diagnosis for evidence of infection. Another advantage of this commercial sVNT is that it allows standardization between clinical laboratories without the need to use live biological materials or biosafety containment. Together with the international unit (IU) recently established by WHO and National Institute of Biological Standards and Controls (NIBSC)(15), this platform could be particularly useful for vaccine evaluation in sub-Saharan Africa, where diagnostic labs lack the infrastructure to run cVNTs. During serosurveillance studies, the test can also be run as an independent test to exclude cross-reactivity after the initial screening with a Luminex MIA assay. Furthermore, given that the test is both species and isotype independent, it could be used as a primary screening assay to detect reversed spillover or spillback events of SARS-CoV-2 from infected humans to wildlife populations or to find the natural reservoir of closely related sarbecoviruses in bats or other animal populations(8,9).

### Ethics statement

Ethical approval to sample from Belgian COVID-19 cases was given by the institutional review boards of the University of Ghent, the University Hospital Antwerp and Jessa Hospital Hasselt. Ethical clearance to use samples for the evaluation of the test was given by the Institutional Review board of the Institute of Tropical Medicine Antwerp. For the Congolese samples, ethical approval was given by Institutional Review board of the Institute of Tropical Medicine Antwerp and the National Reference Laboratory INRB in Kinshasa (DRC). All participants provided informed consent to participate. We declare that the planning conduct and reporting of the study was in line with the Declaration of Helsinki, as revised in 2013.

## Funding

The work was funded by a European & Developing Countries Clinical Trials Partnership (EDCTP) project (Africover: RIA2020EF-3031), the Research Foundation Flanders (FWO) (G0G4220N and G054820N), the Health Care Worker seroprevalence study (Sciensano/ITM), NCT04373889 and intramural funds from the Institute of Tropical Medicine Antwerp. Joachim Marien is currently a research assistant of Research Foundation Flanders (FWO) and Antoine Nkuba received a doctoral scholarship from the French Institut de Recherche pour le Developpement. Work at Duke-NUS is supported by Singapore National Research Foundation (NRF2016NRF-NSFC002-013) and National Medical Research Council (STPRG-FY19-001 and COVID19RF-003). The Congo study was conducted with funding from Enabel (the Belgian Development agency), GIZ (Deutsche Gesellschaft fur Internationale Zusammenarbeit) and the framework agreement between the Institute of Tropical Medicine and the Belgian Development Cooperation.

## Data Availability

The datasets used and/or analysed during the current study are available from the corresponding or last author on reasonable request.

## Author statement

Conceived the study: JMa, L-FW and KA. Wrote the paper: JMa and KA. Performed the lab experiments: JMa, JMi, AN, LH, AC. Performed the statistical analyses: JMa. Supervised data collection and laboratory work: KB, PM, JMad, VvL, SA, L-FW and KA. All authors read and approved the final manuscript.

## Declaration of Competing Interest

L-FW is a co-inventor on a patent application for the sVNT technology and a commercial kit, cPassTM, is being marketed by GenScript Biotech. Other authors declare that they have no known competing financial interests or personal relationships that could have appeared to influence the work reported in this paper.

## Notes

### Clinical Trial

NA

## References

1. Arora RK, Joseph A, Wyk J Van, Rocco S, Atmaja A, May E, et al. SeroTracker: a global SARS-CoV-2 seroprevalence dashboard. Lancet Infect Dis. 2020;(January):19–21.

2. Dan JM, Mateus J, Kato Y, Hastie KM, Faliti CE, Ramirez SI, et al. Immunological memory to SARS-CoV-2 assessed for greater than six months after infection. bioRxiv. 2020;4063(January):1–23.

3. Duysburgh E, Mortgat L, Barbezange C, Dierick K, Fischer N, Heyndrickx L, et al. Persistence of IgG response to SARS-CoV-2. Lancet Infect Dis [Internet]. 2021;21(2):163–4. Available from: http://dx.doi.org/10.1016/S1473-3099(20)30943-9

4. Selhorst P, Ierssel S Van, Michiels J, Mariën J, Bartholomeeusen K, Dirinck E, et al. Symptomatic SARS-CoV-2 re-infection of a health care worker in a Belgian nosocomial outbreak despite primary neutralizing antibody response. Clin Infect Dis. 2020;1–18.

5. Thomson EC, Rosen LE, Shepherd JG, Spreafico R, Filipe S, Wojcechowskyj JA, et al. The circulating SARS-CoV-2 spike variant N439K maintains fitness while evading antibody-mediated immunity. 2020;

6. Mariën J, Ceulemans A, Michiels J, Heyndrickx L, Kerkhof K, Foque N, et al. Evaluating SARS-CoV-2 spike and nucleocapsid proteins as targets for antibody detection in severe and mild COVID-19 cases using a Luminex bead-based assay. J Virol Methods. 2021;288.

7. Yue F, Lidenge SJ, Peña PB, Clegg AA, Wood C. High prevalence of pre-existing serological cross-reactivity against severe acute respiratory syndrome coronavirus-2 (SARS-CoV-2) in sub-Saharan Africa. Int J Infect Dis. 2020;102(January):577–83.

8. Perera RAPM, Ko R, Y. OTT, Hui DSC, Kwan MYM, Brackman CJ, et al. Evaluation of a SARS-CoV-2 Surrogate Virus Neutralization Test for Detection of Antibody in Human, Canine, Cat, and Hamster Sera. J Clin Microbiol. 2021;59(2):1–6.

9. Wacharapluesadee S, Tan CW, Maneeorn P, Duengkae P, Zhu F, Joyjinda Y, et al. Evidence for SARS-CoV-2 related coronaviruses circulating in bats and pangolins in Southeast Asia. Nat Commun [Internet]. 2021;12(1):972. Available from: http://www.ncbi.nlm.nih.gov/pubmed/33563978

10. Tan CW, Chia WN, Qin X, Liu P, Chen MIC, Tiu C, et al. A SARS-CoV-2 surrogate virus neutralization test based on antibody-mediated blockage of ACE2–spike protein– protein interaction. Nat Biotechnol [Internet]. 2020;38(9):1073–8. Available from: http://dx.doi.org/10.1038/s41587-020-0631-z

11. Mortgat L, Barbezange C, Fischer N, Heyndrickx L, Hutse V, Thomas I, et al. SARS-CoV-2 Prevalence and Seroprevalence among Healthcare Workers in Belgian Hospitals: Baseline Results of a Prospective Cohort Study Authors*. medRxiv [Internet]. 2020;2020.10.03.20204545. Available from: https://doi.org/10.1101/2020.10.03.20204545

12. Ndaye AN, Hoxha A, Madinga J, Mariën J, Peeters M, Leendertz FH, et al. Challenges in interpreting SARS-CoV-2 serological results in African countries. Lancet Glob Heal. 2021;(21):19–20.

13. Bond K, Nicholson S, Lim SM, Karapanagiotidis T, Williams E, Johnson D, et al. Evaluation of serological tests for SARS-CoV-2: Implications for serology testing in a low-prevalence setting. J Infect Dis. 2020;222(8):1280–8.

14. Meyer B, Reimerink J, Torriani G, Brouwer F, Godeke GJ, Yerly S, et al. Validation and clinical evaluation of a SARS-CoV-2 surrogate virus neutralisation test (sVNT). Emerg Microbes Infect. 2020;9(1):2394–403.

15. Mattiuzzo G, Bentley EM, Hassall M, Routley S. Establishment of the WHO International Standard and Reference Panel for anti-SARS-CoV-2 antibody. World Heal Organ [Internet]. 2020;(December):9–10. Available from: https://www.who.int/publications/m/item/WHO-BS-2020.2403

